# Automatic Classification of Medical Artificial Intelligence Articles by Their Level of Translational Maturity: An Interpretable Supervised Text-Classification Approach

**DOI:** 10.64898/2026.07.09.26357253

**Authors:** Sandeep Reddy, Alix Héritier

## Abstract

The rapid expansion of the medical artificial intelligence (AI) literature has outpaced our ability to judge how far published models have progressed towards clinical use. We investigated whether the translational maturity of a study can be estimated automatically from its abstract. Using PubMed, we assembled a corpus of 11,024 candidate articles, reduced it to 1,816 AI-related articles by heuristic filtering, and manually double-annotated a balanced sample of 524 articles across five maturity classes (internal validation, external validation, prospective evaluation, implementation or governance, and not applicable). Abstracts were represented as TF-IDF features and classified using multinomial logistic regression with a Lasso penalty, chosen for interpretability and suitability for a small, imbalanced dataset. On a stratified held-out test set (n = 104), the model achieved 69.2% accuracy, Cohen’s kappa of 0.495, macro-F1 of 0.458 and a weighted AUC of 0.820. Performance was strong for the frequent classes but poor for the rare implementation or governance class, which the model failed to recover. A balanced manual verification of 200 large-corpus predictions confirmed this pattern, with per-class precision ranging from 82.5% (internal validation) to 5.0% (implementation or governance). An interpretable, low-resource classifier can support literature mapping but requires human oversight for advanced maturity levels.

## 1. Introduction

Artificial intelligence is increasingly prominent in medical research. Many published articles present models intended to support the analysis of medical images, the detection of pathologies, risk prediction, or stages of clinical decision-making [Esteva et al., 2019]. This substantial output reflects strong momentum in the field, but it also makes it more difficult to assess the overall progress achieved by the published work. As the volume of publications increases, manual review alone becomes less practical for identifying which studies remain at an early modelling stage and which have advanced towards clinical evaluation. A scalable method for organising this literature could therefore help researchers, clinicians, and policymakers distinguish technical development from evidence that is closer to real-world translation.

Not all articles on medical AI are equally mature. Some studies are limited to model development and evaluation on internal data, while others go further, conducting external validation, prospective evaluation, or a study of integration into a real clinical setting [Kelly et al., 2019; Nagendran et al., 2020]. This distinction matters because a model that performs well in an experimental setting cannot be considered ready for clinical practice. The capacity of a model to generalise to other populations, institutions, or real-world conditions of use is a central issue for translation [Subbaswamy & Saria, 2020; Zech et al., 2018].

In this context, we studied the extent to which scientific articles can be automatically classified by their level of translational maturity using information available in their abstracts. The task can be framed as follows: *how can the reading and sorting of scientific articles be transformed into a supervised classification problem that helps map the literature on medical AI?* The units of study are scientific articles, described mainly by an abstract, a PubMed identifier, a year of publication, and a manually assigned maturity label. The approach relies on an automated method for analysing scientific text. Abstracts are transformed into numerical variables using TF-IDF, quantifying the relative importance of words or word groups in each document [Salton & Buckley, 1988]. These variables are then used in a multinomial logistic regression with a Lasso penalty [Friedman et al., 2010]. This model was chosen because it is well-suited to multi-class classification while remaining readable: it produces predictions and also identifies the terms associated with each class.

### Contributions

This study makes three contributions. First, we define an operational five-class taxonomy of translational maturity for medical AI articles and construct a double-annotated reference dataset of 524 articles. Second, we develop a fully reproducible, interpretable classification pipeline and evaluate it on a stratified held-out test set. Third, we apply the model to a larger unannotated corpus to construct an exploratory evidence map, and we validate a balanced sample of these predictions by manual review, quantifying where automated classification is and is not reliable.

## 2. Methods

### 2.1 Study design and data source

Data were drawn from the scientific literature available on PubMed, a bibliographic database widely used in the biomedical field that provides references, metadata, and abstracts. To identify relevant articles, a search filter was constructed to select medical AI articles likely to fall within the study’s scope. Applying this filter to PubMed yielded an initial corpus of 11,024 articles. This corpus was deliberately broad to limit premature exclusion of relevant work. Of the 11,024 articles, 11,007 (99.8%) had an abstract; only 17 lacked one. The PubMed database was chosen because it is an open-access resource that supports reproducibility without an institutional subscription.

A second heuristic filter was applied in R to retain articles that presented sufficient textual cues of an AI link, based on the presence of terms such as machine learning, deep learning, and neural networks. This filter was not intended as a definitive annotation but to reduce the corpus to a more targeted set. Following it, the corpus was reduced from 11,024 to 1,816 articles, forming the unannotated corpus from which a reference dataset was built.

### 2.1a Search strategy

The search was executed in PubMed on 4^th^ May 2026 and combined artificial-intelligence terms with a medical or clinical scope. The search was restricted to English-language research articles involving humans, published from 2018 onward, with reviews, editorials, conference abstracts, and protocols excluded. This deliberately broad query prioritised recall over precision, with specificity recovered during subsequent filtering and annotation rather than in the query itself. The full search strategy is outlined in the Appendix.

This query returned 11,024 records. Of these, 11,007 (99.8%) carried an abstract, and 17 did not. A second heuristic filter was then applied in R to retain records showing sufficient lexical evidence of an AI or machine-learning method, based on the presence of at least one predefined term (for example, machine learning, deep learning, or neural networks) in the title or abstract. This lexical screen, which was not a human relevance judgement, reduced the corpus from 11,024 to 1,816 records, forming the unannotated corpus from which the reference dataset was built.

### 2.1b Inclusion and exclusion criteria

A record was included if it satisfied all of the following: was indexed in PubMed; was returned by the search query above; had a retrievable abstract; and contained at least one AI or machine-learning term from the predefined list. A record was excluded if any of the following held: it had no retrievable abstract (17 records); it failed the heuristic AI-term screen (insufficient lexical evidence of an AI or machine-learning method); it was not an original empirical study of an evaluable AI model / was not applicable to the translational-maturity taxonomy, or it carried a duplicate PubMed identifier.

The publication window of 2018 to 4 May 2026 and the balancing across publication years were applied at the annotation-sampling stage rather than at the search stage; the corpus was not year-restricted at source. The not-applicable category (class 5) is an annotation label retained in the dataset, not a pre-annotation exclusion criterion: reviews, editorials, and conceptual articles that passed both filters were retained and labelled class 5 rather than removed.

### 2.1c Data extraction sheet

The following fields were extracted for each record. The first block comprises metadata drawn from PubMed; the second is assigned during annotation and reconciliation.

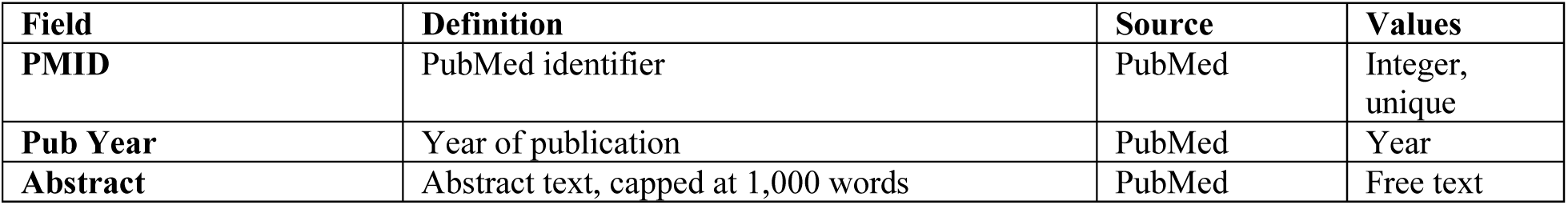
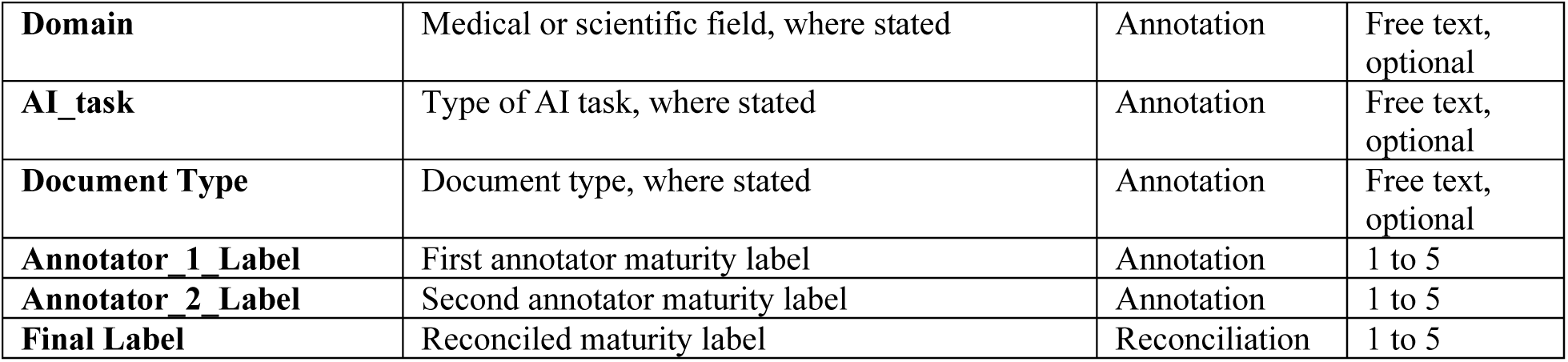

### 2.1d Corpus assembly flowchart

**Figure 1** summarises the corpus assembly.

**Figure 1.**
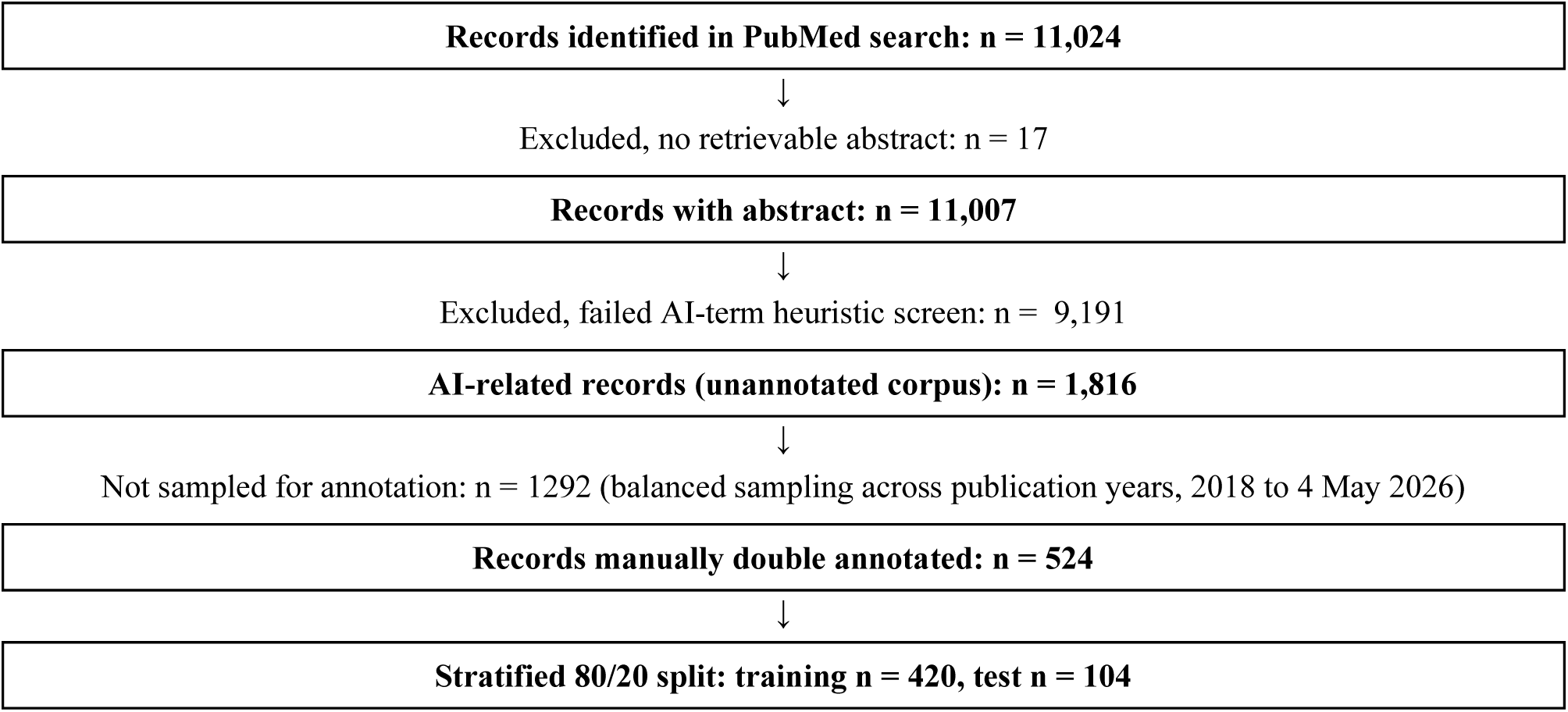
Flowchart of corpus assembly and inclusion/exclusion decisions.

### 2.2 Construction of the annotated dataset

The 1,816 filtered articles carried no maturity label, so a subset was selected for manual annotation. A sample was drawn in R from articles published between 2018 and the start of the study (4 May 2026), balanced across publication years to prevent the most recent years from dominating. This yielded 524 articles for manual annotation, each described by an abstract, a PubMed identifier, the year of publication, and, after annotation, a maturity label. Abstracts corresponded to those available on PubMed, with a maximum length of 1,000 words.

The statistical unit is the scientific article. The main variables used in the model were the article identifier, publication year, abstract, and maturity label. Additional descriptive variables (application field, AI task type, document type) were retained for corpus characterisation and error analysis but were not used as model inputs. **Table 1** summarises these variables.

**Table 1.** Main variables used in the study.

| Variable | Description | Role in the study |
| --- | --- | --- |
| **PMID** | PubMed identifier of the article | Unique identification of articles |
| **Pub Year** | Year of publication | Temporal description of the corpus |
| **Abstract** | Scientific abstract available on PubMed, limited to a maximum of 1,000 words | Textual information used for classification |
| **Domain** | Maturity level assigned to the article | Target variable of the model |
| **AI_task** | Medical or scientific field of the article, where available | Additional descriptive variable |
| **Document_Type** | Type of artificial intelligence task, where available | Additional descriptive variable |
| **Annotator_1_Label** | Type of document, where available | Additional descriptive variable |

The taxonomy distinguished four levels of translational maturity, with a fifth class added for articles that cannot be placed on this scale (for example, systematic reviews, commentaries, conceptual articles, or work without empirical evaluation of an AI model). The five classes are defined in **Table 2**.

**Table 2.** Definition of the five maturity classes.

| **Label** | **Interpretation** |
| --- | --- |
| **1** | Internal validation: the model is developed and evaluated on data from the same general source as the training data, for example using a train-test split, cross-validation or resampling. |
| **2** | External validation: the model is evaluated on data independent of the development set, for example from another institution, population, period or external cohort. |
| **3** | Prospective evaluation: the model is evaluated on prospectively collected data, in a real or simulated clinical pathway setting, with an evaluation protocol defined in advance. |
| **4** | Implementation or governance: the study describes an operational use, a deployment, follow-up, a monitoring process, a clinical integration or governance elements associated with the model. |
| **5** | Not applicable: the article cannot be classified on the maturity scale (for example a review, editorial, commentary, or methodological article without empirical validation). |

Class 2 was assigned only when the abstract explicitly mentioned validation on independent data; the mere presence of multicentre data was insufficient. This strict rule reduces overestimation of maturity but makes annotation more demanding.

### 2.3 Annotation procedure and reliability

To ensure annotation quality, the 524 articles were double-annotated: first and second. The two label sets were compared to assess inter-annotator agreement, using Cohen’s kappa, which quantifies observed agreement while accounting for chance. Disagreements were examined in a reconciliation step to assign a final label to each article, limiting individual errors and strengthening dataset consistency. The resulting class distribution is given in **Table 3**.

**Table 3.** Distribution of the annotated dataset by class.

| **Label** | **Number of articles** | **Proportion** |
| --- | --- | --- |
| **1** | 280 | 53.4% |
| **2** | 142 | 27.1% |
| **3** | 36 | 6.9% |
| **4** | 6 | 1.1% |
| **5** | 60 | 11.5% |
| **Total** | 524 | 100.0% |

Classes 1 and 2 dominate, whereas classes 3 and especially 4 are sparse. This imbalance is important for interpreting the results because a supervised model learns classes with more observations more easily.

### 2.4 Data preparation and train-test split

Several checks preceded modelling. Missing values were limited: 11,007 of 11,024 articles had an abstract, and articles without an abstract or a label were excluded from steps that required that information. Label consistency was verified against the five defined classes. The annotated corpus was then split into training and test sets in a stratified manner to preserve, as far as possible, the representation of each class. **Table 4** reports the resulting distribution.

**Table 4.**
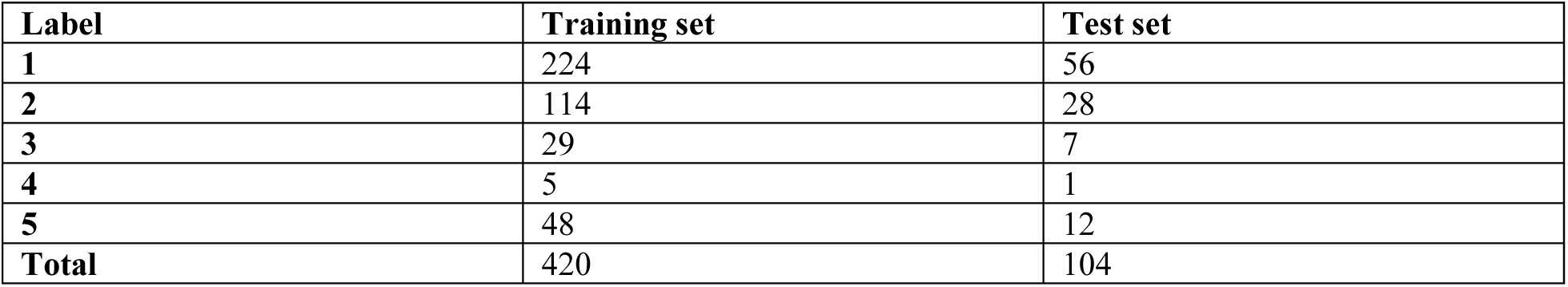
Distribution of the classes in the training and test sets (80/20 stratified split).

Stratification is critical when class imbalance is present: without it, rare classes could be absent from the test set, rendering the evaluation uninformative. Class 4 nevertheless remains very weakly represented, with five training and one test article, making its evaluation fragile.

### 2.5 Feature representation

Abstracts were transformed into a vector of numerical explanatory variables using TF-IDF, which weights words and word groups by their relative importance in each document. Each abstract is therefore represented by a high-dimensional numerical vector, and these variables are used to estimate the probability that an article belongs to each of the five classes. Descriptive analysis of abstract length (**Figure 2**) informed this representation: very short abstracts may contain too little information for reliable classification, whereas more detailed abstracts may explicitly mention the type of validation performed.

**Figure 2.**
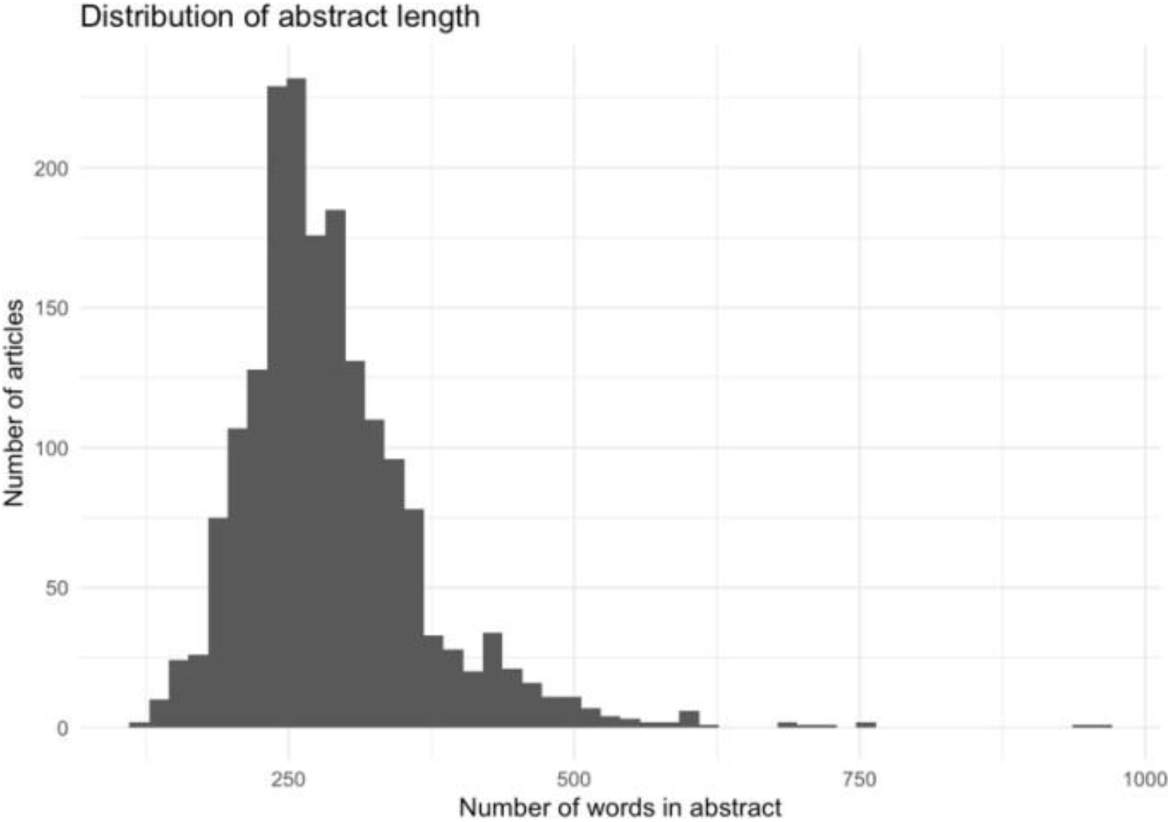
Distribution of abstract length across the corpus.

### 2.6 Statistical model

Each observation corresponds to an article identified by a unique PubMed identifier. The variable to be explained is the maturity label Yᵢ, taking values in:

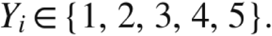

corresponding respectively to internal validation, external validation, prospective evaluation, implementation or governance, and the not applicable class. The problem is therefore a multi-class supervised classification problem. The abstract of each article is transformed into a vector of numerical explanatory variables:

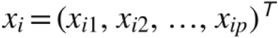

where p denotes the number of textual variables retained after preprocessing, corresponding to the TF-IDF weights of the terms present in the abstracts.

For context, in binary logistic regression, the response takes two values, Yᵢ ∈ {0, 1}, and we write:

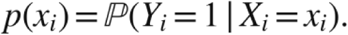

The probability is assumed to be linearly related to the explanatory variables through the logit function:

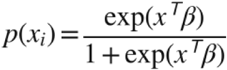

equivalently:

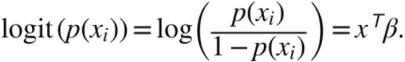

This means that it is the log-ratio of the probability of belonging to class 1 to that of not belonging, rather than the probability itself, that is assumed linear in the explanatory variables. Because the target has five categories, we use multinomial logistic regression:

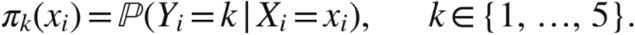

With class 1 as a reference, the model compares each class k to it. For k ∈ {2, 3, 4, 5}:

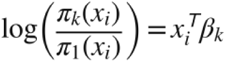

Each class therefore has its own coefficient vector βₖ. A positive coefficient for a term increases the probability of belonging to class k relative to the reference class; a negative coefficient decreases it. Predicted probabilities are obtained using the softmax function. For the reference class:

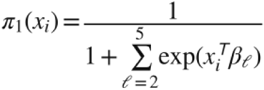

and for a class k ∈ {2, 3, 4, 5}:

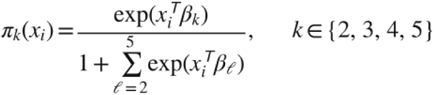

The model assigns to the article the class with the highest predicted probability:

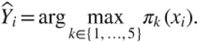

Estimation relies on the multinomial likelihood. Introducing the indicator yᵢₖ, equal to 1 if article i belongs to class k and 0 otherwise, the log-likelihood is:

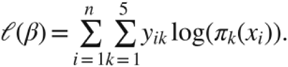

For textual data, the number of explanatory variables can be very large relative to the number of annotated articles, risking an over-complex model that generalises poorly. A Lasso penalty is therefore added, and estimation minimises the negative penalised log-likelihood:

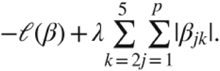

The parameter λ controls the penalty strength. Small λ retains more non-zero coefficients; large λ shrinks more coefficients to zero. This enables automatic selection of the terms most useful for classification. The model assumes independence among observations (each row is a distinct article with a unique identifier), linearity on the logit scale in the TF-IDF weights, and the absence of perfect separation, which is mitigated by the Lasso. As a generalised linear model for a qualitative response, it does not assume normality or homoscedasticity of errors.

### 2.7 Model selection, choice, and validation

A multinomial logistic regression with a Lasso penalty was selected under four constraints. The target is a qualitative five-category variable, so linear regression is unsuitable. The annotated corpus is small (524 articles) and includes underrepresented classes, so an overcomplex model would overfit. The TF-IDF representation is high-dimensional, which the Lasso addresses by shrinking coefficients to zero. Finally, interpretability was required, and penalised logistic regression exposes the terms associated with each class. Simpler baselines were rejected: a majority-class predictor would be useless in practice, and keyword rules would miss paraphrases and misfire in ambiguous contexts (for example, an abstract mentioning future implementation). More complex biomedical language models were considered but were not selected because they generally impose greater computational demands, require a substantial amount and diversity of training data, and offer less direct feature-level interpretability than sparse linear models [Lee et al., 2020; Rudin, 2019].

Articles lacking a label, PubMed identifier, publication year, or abstract were removed, and labels were restricted to classes 1-5. An 80/20 stratified split by label was applied (**Table 4** above; 420 training, 104 test articles). The penalty parameter λ was selected within the training set over a grid of candidate values, each evaluated on a validation portion of the training data, so the test set was never used for selection. The primary selection criterion was Cohen’s kappa, more informative than accuracy under imbalance, with accuracy as a complementary indicator. Once λ was selected, the final model was retrained on the full training set and applied to the test set. The workflow was implemented as a reproducible R pipeline (data preparation, split, TF-IDF transformation, penalised multinomial training, evaluation) producing a confusion matrix, global measures, ROC curves, calibration analysis, and coefficient extraction.

## 3. Results

### 3.1 Model performance on the held-out test set

The model was validated on the test set of 104 articles not used during training. Global indicators are reported in **Table 5**.

**Table 5.** Global performance indicators on the test set.

| Metric | Value |
| --- | --- |
| Accuracy | 69.2% |
| Cohen's kappa | 0.495 |
| Macro-F1 | 0.458 |
| Weighted AUC | 0.820 |
| Multinomial log-loss | 1.27 |

Accuracy reached 69.2%, meaning about seven articles in ten were correctly classified. Cohen’s kappa of 0.495 indicates moderate agreement after adjusting for chance. The macro-F1 of 0.458 reflects heterogeneous performance across classes, particularly for the least-represented ones. The weighted AUC of 0.820 indicates that the model discriminates between classes based on predicted probabilities, while the log-loss of 1.27 shows that some predicted probabilities remain uncertain, especially for rare classes.

The confusion matrix (**Figure 3**) shows that 72 of 104 test articles were correctly classified. Class 1 was recognised particularly well (44 of 56 correct), and the majority of class 2 articles were identified (21 of 28). Errors were consistent with the proximity between internal and external validation: several class 1 articles were predicted as class 2 or class 5, and some class 2 articles as class 1. Class 3 was harder to identify (2 of 7 correct), and class 4 was not recovered, as anticipated from its single test article and five training articles.

**Figure 3.**
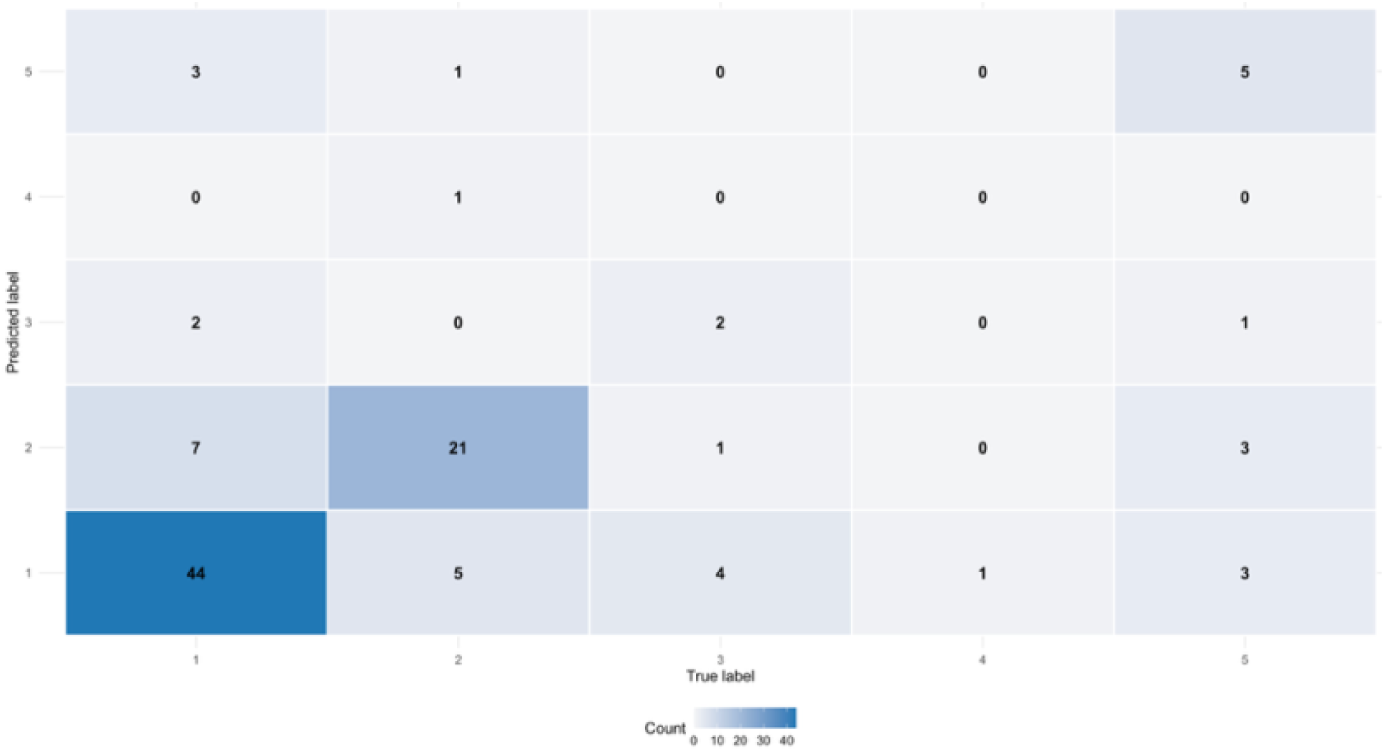
Confusion matrix on the test set (true label against predicted label).

One-versus-rest ROC curves (**Figure 4**) gave an overall weighted AUC of 0.820. Classes 1, 2 and 3 showed high AUCs of 0.835, 0.825 and 0.882, respectively, indicating good ranking of articles by class probability. Class 5 showed a more moderate AUC of 0.762, whereas class 4 showed a very low AUC of 0.136, indicating that the model failed to discriminate between these classes.

**Figure 4.**
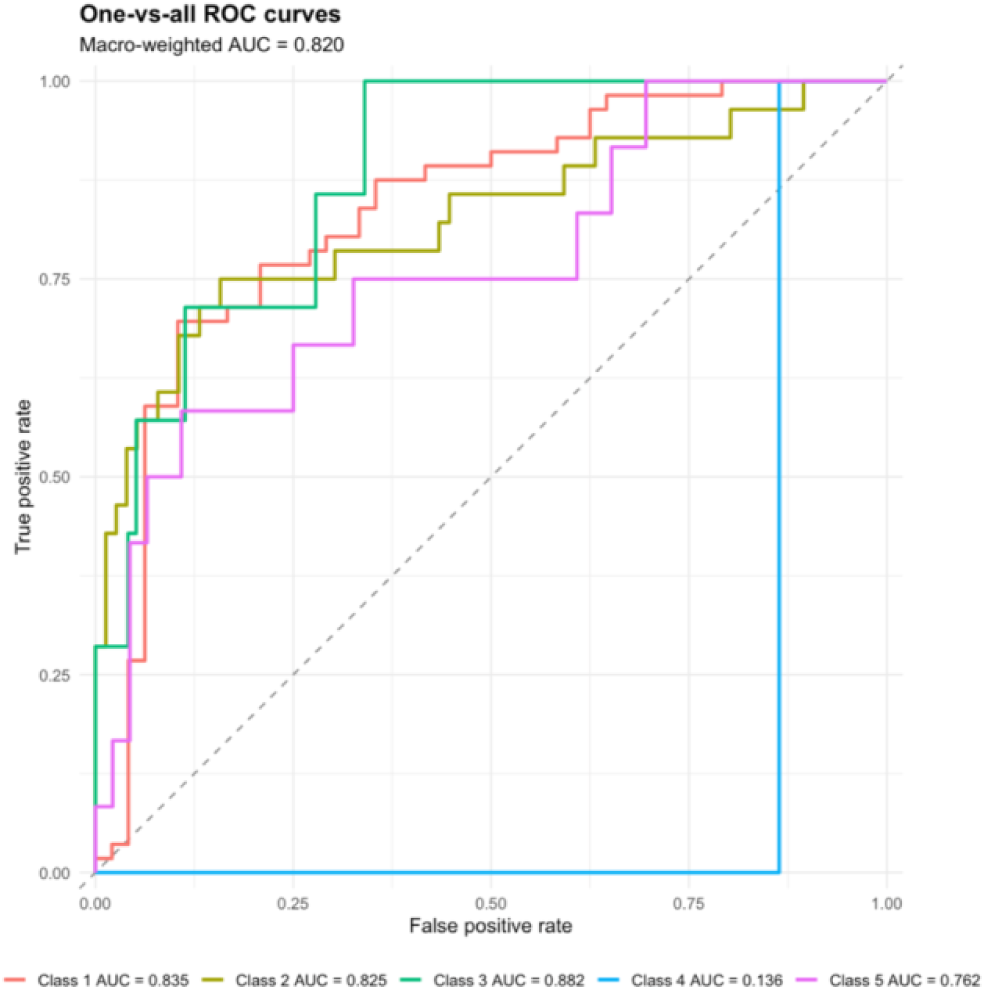
ROC curves by class (one class against all the others).

Calibration analysis (**Figure 5**) assessed whether predicted probabilities matched observed frequencies. Calibration was relatively usable for classes 1 and 2, which have more observations, but much more fragile for classes 3, 4 and 5. Class 4 is almost impossible to interpret because it is underrepresented, and for class 5, the curve generally lies below the diagonal, suggesting probabilities that are too high relative to the observed frequency.

**Figure 5.**
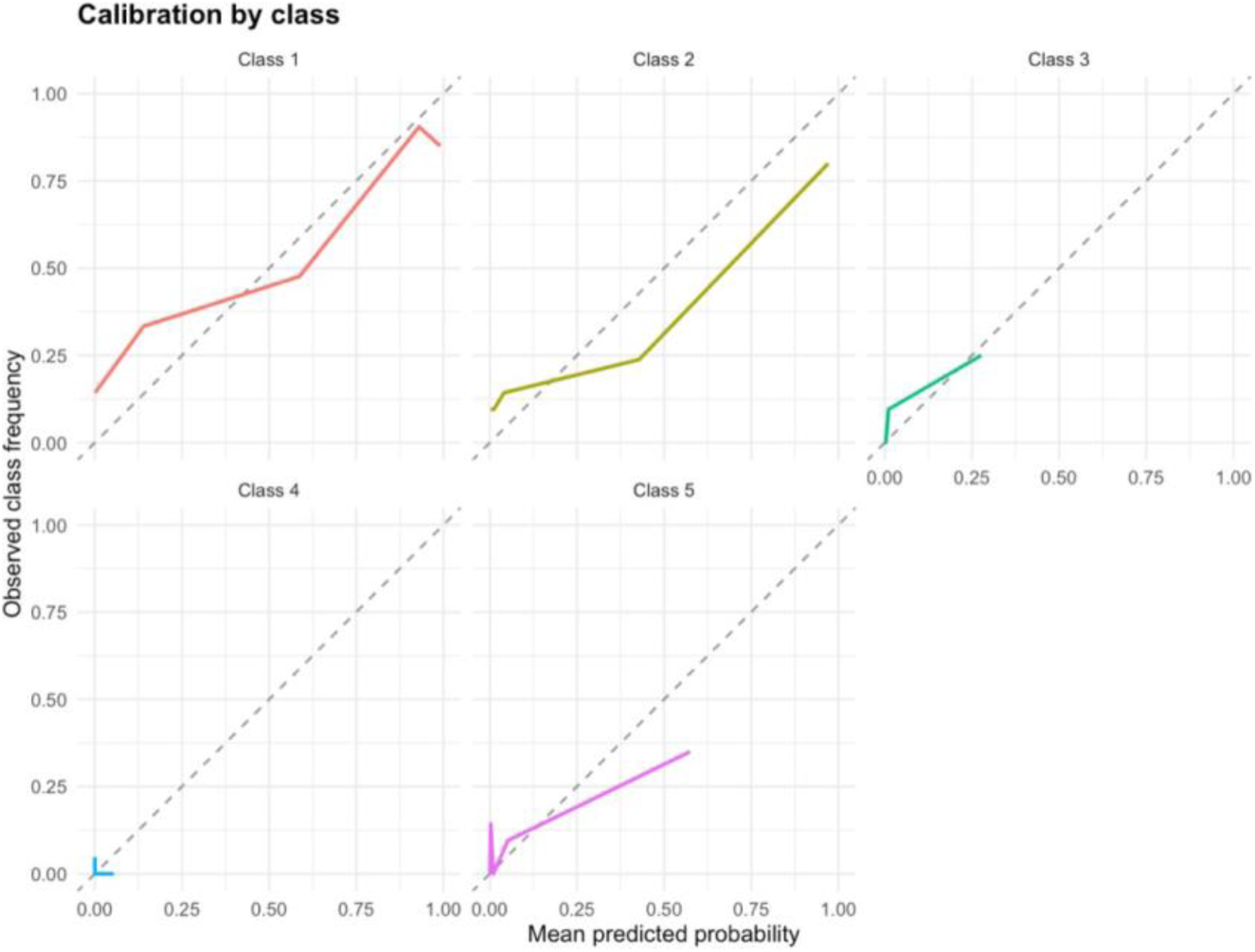
Calibration of the predicted probabilities by class.

### 3.2 Application to the large corpus and evidence map

After evaluation, the model was applied to the larger unannotated corpus. For each article, the model produced a probability for each of the five classes and assigned a predicted class, enabling estimation of the distribution of maturity levels across a set far larger than the annotated dataset (**Figure 6**) and cross-referencing with publication year to examine temporal evolution (**Figure 7**).

**Figure 6.**
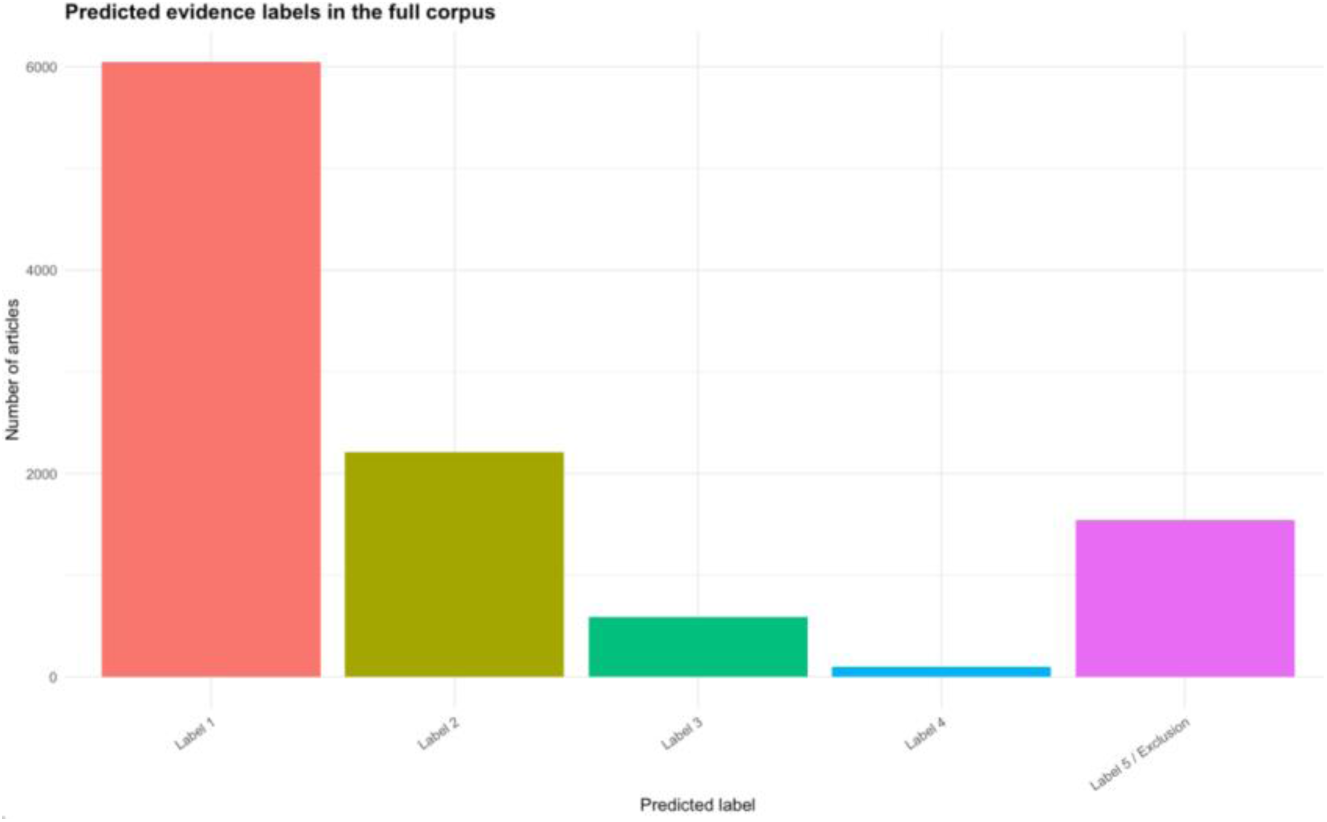
Predicted evidence labels in the full corpus.

**Figure 7.**
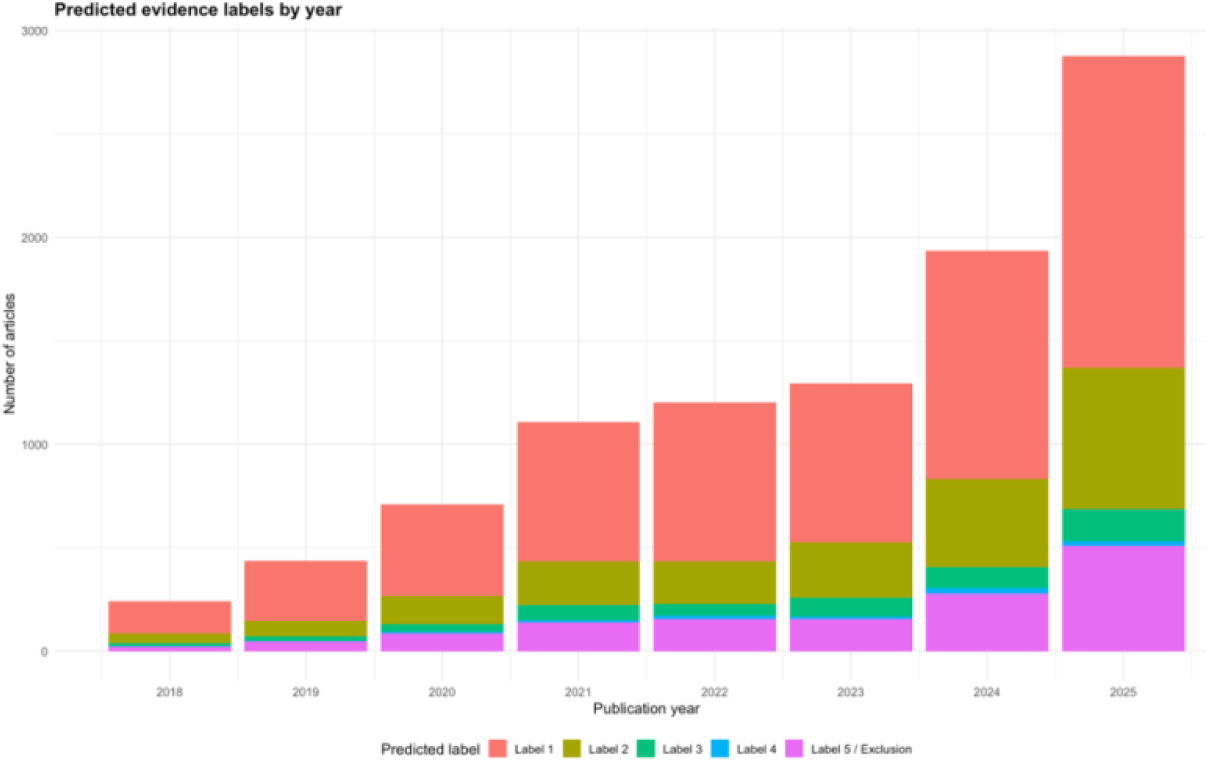
Predicted evidence labels by year of publication.

This step transforms the statistical model into a tool that supports reading the literature, giving an overview of whether research is progressing towards more robust validations, whether studies move beyond internal validation, and whether real clinical integration remains marginal. The mapping must be interpreted with caution: the predicted classes are model outputs from a limited annotated dataset, not definitive human annotations, and the evidence map is therefore an exploratory instrument.

### 3.3 Manual verification of large-corpus predictions

To evaluate prediction quality on the large corpus, a balanced sample of 200 predicted articles was constituted, 40 per predicted class, enabling class-by-class assessment including rare classes. These articles were double-annotated, compared using Cohen’s kappa, and reconciled to a final reference label. Because the sample is balanced rather than random, the metrics evaluate per-class reliability rather than overall corpus performance.

Of the 200 verified articles, 97 were correctly classified (accuracy 48.5%), with Cohen’s kappa of 0.356 and macro-F1 of 0.446 against the reconciled labels (**Figure 8**). Per-class precision (**Table 6**) was highest for class 1 (33 of 40 confirmed; 82.5%) and solid for class 2 (27 of 40; 67.5%), but fragile for classes 3, 4 and 5.

**Figure 8.**
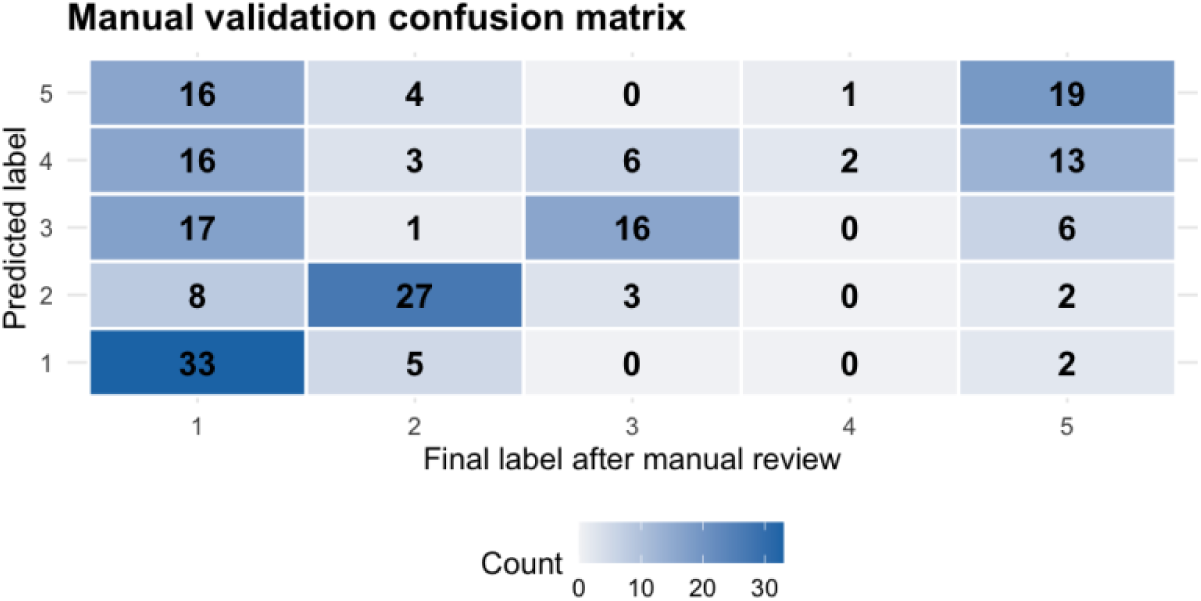
Comparison matrix between predicted labels and final labels after manual review.

**Table 6.** Per-class precision of the predictions on the manually reviewed sample.

| Predicted class | Articles predicted | Correctly confirmed | Precision |
| --- | --- | --- | --- |
| 1 | 40 | 33 | 82.5% |
| 2 | 40 | 27 | 67.5% |
| 3 | 40 | 16 | 40.0% |
| 4 | 40 | 2 | 5.0% |
| 5 | 40 | 19 | 47.5% |

Class 4 was especially problematic: of 40 articles predicted as implementation or governance, only 2 were confirmed, most being reclassified as class 1 or class 5. This is consistent with class 4’s rarity in the annotated dataset (six articles total, five in training), which leaves the model unable to distinguish a genuine implementation study from an article merely mentioning potential clinical use. Excluding the 40 class 4 predictions (**Table 7**), accuracy rose from 48.5% to 59.4%, kappa from 0.356 to 0.459, and macro-F1 from 0.446 to 0.593.

**Table 7.**
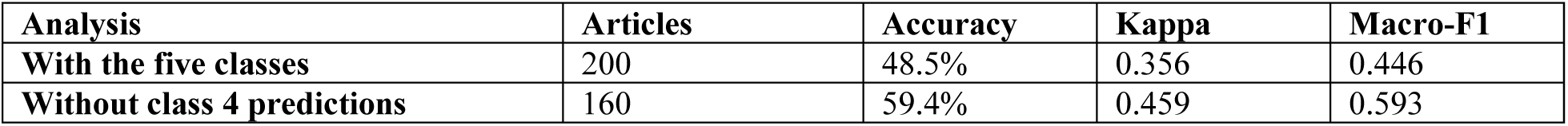
Comparison of the metrics with and without the class 4 predictions.

| Analysis | Articles | Accuracy | Kappa | Macro-F1 |
| --- | --- | --- | --- | --- |
| With the five classes | 200 | 48.5% | 0.356 | 0.446 |
| Without class 4 predictions | 160 | 59.4% | 0.459 | 0.593 |

This confirms that class 4 predictions substantially degrade overall performance, driven by the small number of examples and the class’s conceptual difficulty. For operational use, articles predicted as class 4 should be re-read systematically before being interpreted as genuine implementation studies.

### 3.4 Feature importance

Because the model uses a Lasso penalty, some coefficients are shrunk to zero, and the retained terms can be interpreted as textual cues associated with certain classes. The aim is not to interpret exact coefficient values but to check whether the selected terms are consistent with the maturity definitions (**Table 8**).

**Table 8.** Examples of terms positively associated with each class by the model.

| Class | Examples of terms positively associated by the model |
| --- | --- |
| **1 — Internal validation** | single center, fold cross, retrospectively, convolutional neural, model proposed, accuracy |
| **2 — External validation** | external, datasets across, another hospital, model tested, patient dataset, previously reported |
| **3 — Prospective evaluation** | prospective, prospective study, prospective cohort, prospective multicenter, multicenter study, patients aged |
| **4 — Implementation or governance** | report, feasibility study, loop, sorted |
| **5 — Not applicable** | pubmed, searched, meta, challenges, insufficient, medical |

These terms are broadly consistent with the class definitions: class 1 with single-centre and cross-validation cues, class 2 with external or independent-data cues, and class 3 with prospective-study cues. Interpretation is more fragile for class 4, whose associated terms are fewer and less specific and may appear in varied contexts, reflecting its poor performance in manual verification. Class 5 is associated with terms such as PubMed, searched, and meta, consistent with its role as an exclusion class. A coefficient does not prove causation; it only indicates a statistical association within the annotated corpus, so coefficient analysis complements but does not replace a methodological reading of the articles.

## 4. Discussion

### 4.1 Key findings

An interpretable, low-resource classifier can estimate translational maturity from abstracts with moderate overall agreement (kappa 0.495) and useful discrimination (weighted AUC 0.820), but performance is strongly class-dependent. The frequent classes, internal and external validation, are recovered reliably, whereas the advanced maturity classes, prospective evaluation and especially implementation or governance, are not. The manual verification reinforces this: class 1 predictions were confirmed 82.5% of the time, but only 5.0% of class 4 predictions were genuine.

### 4.2 Implications

From a literature-mapping perspective, the model shifts the task from article-by-article reading to synthesis, identifying which maturity levels dominate and how they evolve over time. False positives in advanced classes are the most sensitive errors because they can suggest that an article is closer to clinical use than it is. The results therefore support using the model as a triage and exploration aid rather than a definitive decision tool [Marshall & Wallace, 2019; van de Schoot et al., 2021], with the class 5 exclusion category preventing articles from being forced onto a scale that does not fit them.

### 4.3 Limitations

Several limitations qualify these results. First, maturity was labelled from abstracts, which may omit the methodological detail needed to determine maturity with certainty; this particularly affects the strict internal-versus-external distinction. Second, class imbalance is severe: class 4 has six articles in total, with one in the test set, so its metrics are highly unstable, as manual verification confirmed. Third, reliance on abstracts rather than full texts omits information that could refine classification. Fourth, the TF-IDF representation captures term frequency but not sentential meaning, so a stated intention to conduct external validation in the future may be mistaken for a completed one. Fifth, residual coding or annotation ambiguities may remain despite checks. Finally, the test set is small (104 articles), so the measured performance is a useful but uncertain estimate, and the large-scale predictions do not replace article-by-article validation.

### 4.4 Future directions

The most direct improvement is to enrich the annotated dataset for the least-represented classes, adding examples of prospective evaluation and implementation to strengthen recognition of advanced maturity. Incorporating open-access full texts, where available, would go beyond the limits of abstracts. Finally, the most uncertain predictions could be routed to a further round of manual annotation, in a logic of progressive, human-in-the-loop refinement, and class 4 predictions in particular could be treated as a category requiring systematic human validation.

## 5. Conclusion

This study shows that the reading and sorting of medical AI articles can be transformed into a supervised classification problem. From a double-annotated dataset, a multinomial logistic regression with a Lasso penalty on TF-IDF features recognised a substantial proportion of articles, particularly for the well-represented internal and external validation classes, and its application to a larger corpus produced a first exploratory evidence map. The approach is nevertheless constrained by the information available in abstracts, class imbalance, and the difficulty of the advanced maturity classes, so its outputs must accompany, not replace, a critical human reading. The model offers a reproducible and interpretable basis for the more systematic mapping of translational maturity in the medical AI literature and of its progression towards real clinical use.

## Data and Code Availability

The annotated dataset, R scripts, and the full reproducible pipeline (data preparation, train-test split, TF-IDF feature construction, penalised multinomial training, and evaluation) are available in the project repository: https://github.com/alix-data/trans-maturity

## Author Contributions

S.R. conceived and supervised the study, defined the translational maturity taxonomy and annotation strategy, and contributed to the second annotation and reconciliation. A.H. designed and implemented the reproducible R pipeline, performed feature construction, model training and evaluation, produced the analyses and figures, and led the first annotation. Both authors annotated the reliability samples, interpreted the results, and approved the final manuscript.

## Data Availability

All data produced in the present work are contained in the manuscript

https://github.com/alix-data/trans-maturity

## Acknowledgements

The authors thank the QUT Centre for Data Science for hosting this work.

## Appendix PubMed Corpus Search Strategy

### 1. Core concept blocks

#### 1.1 Artificial intelligence/machine learning block

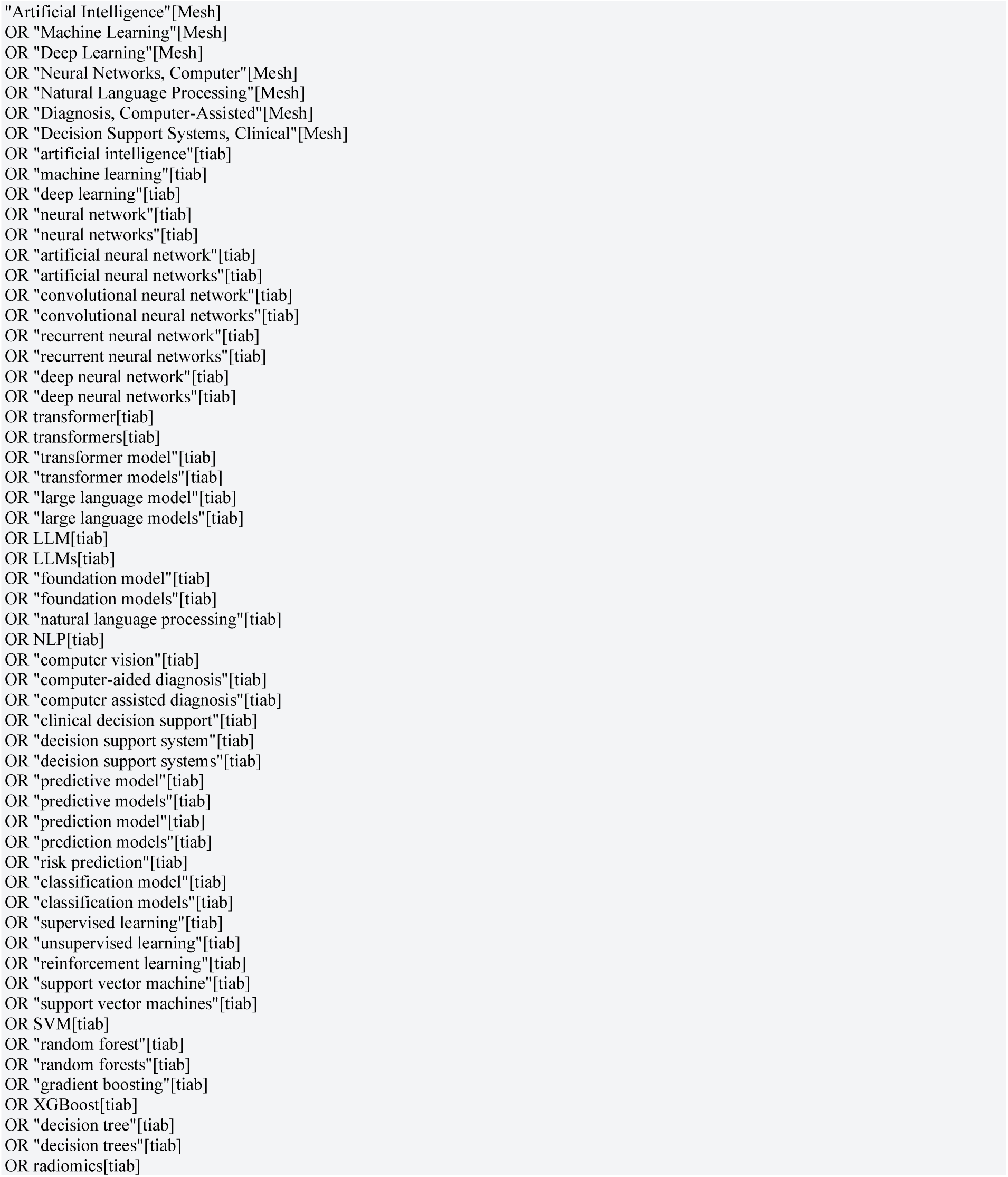

#### 1.2 Medical/clinical scope block

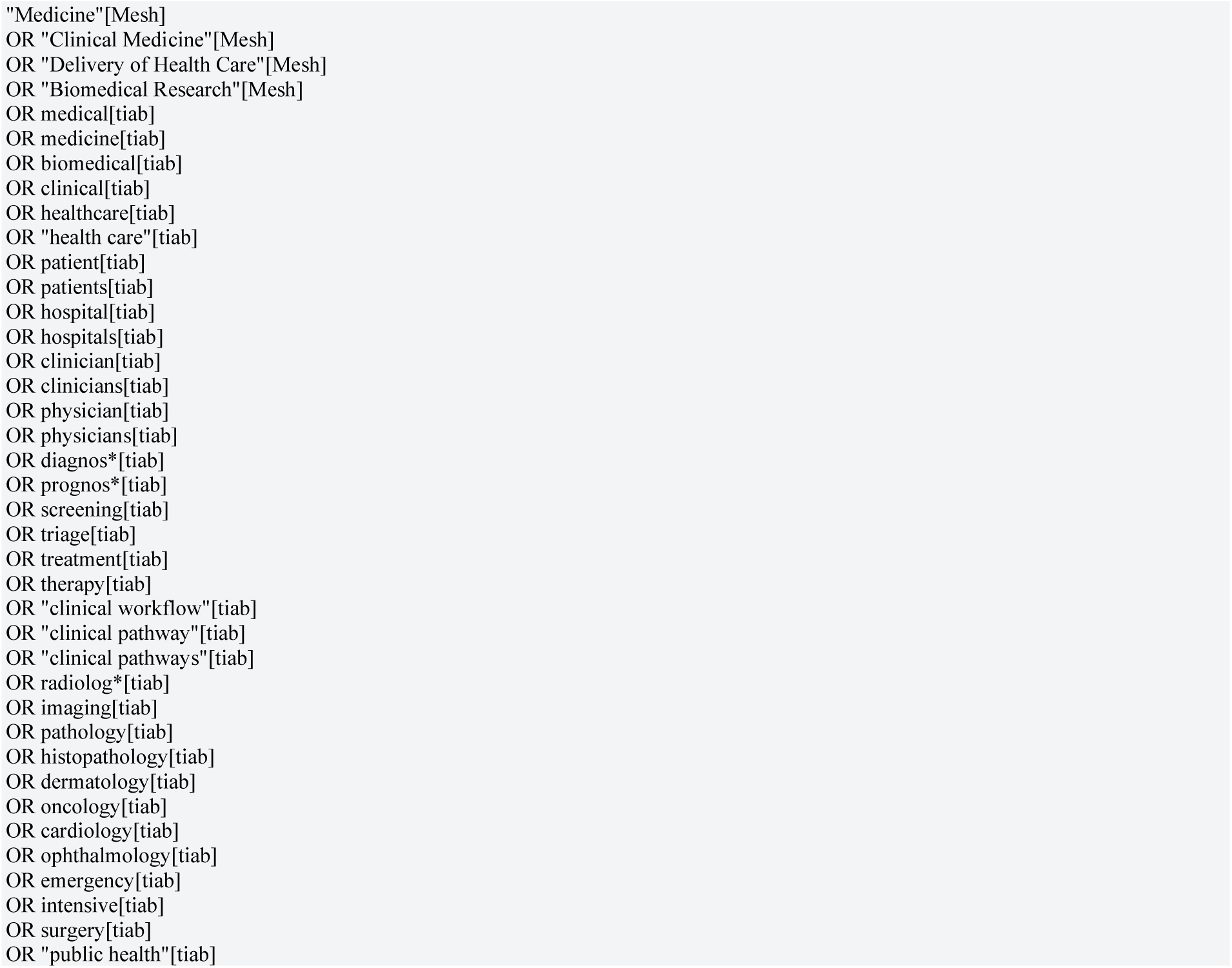

### 2. PubMed broad corpus query

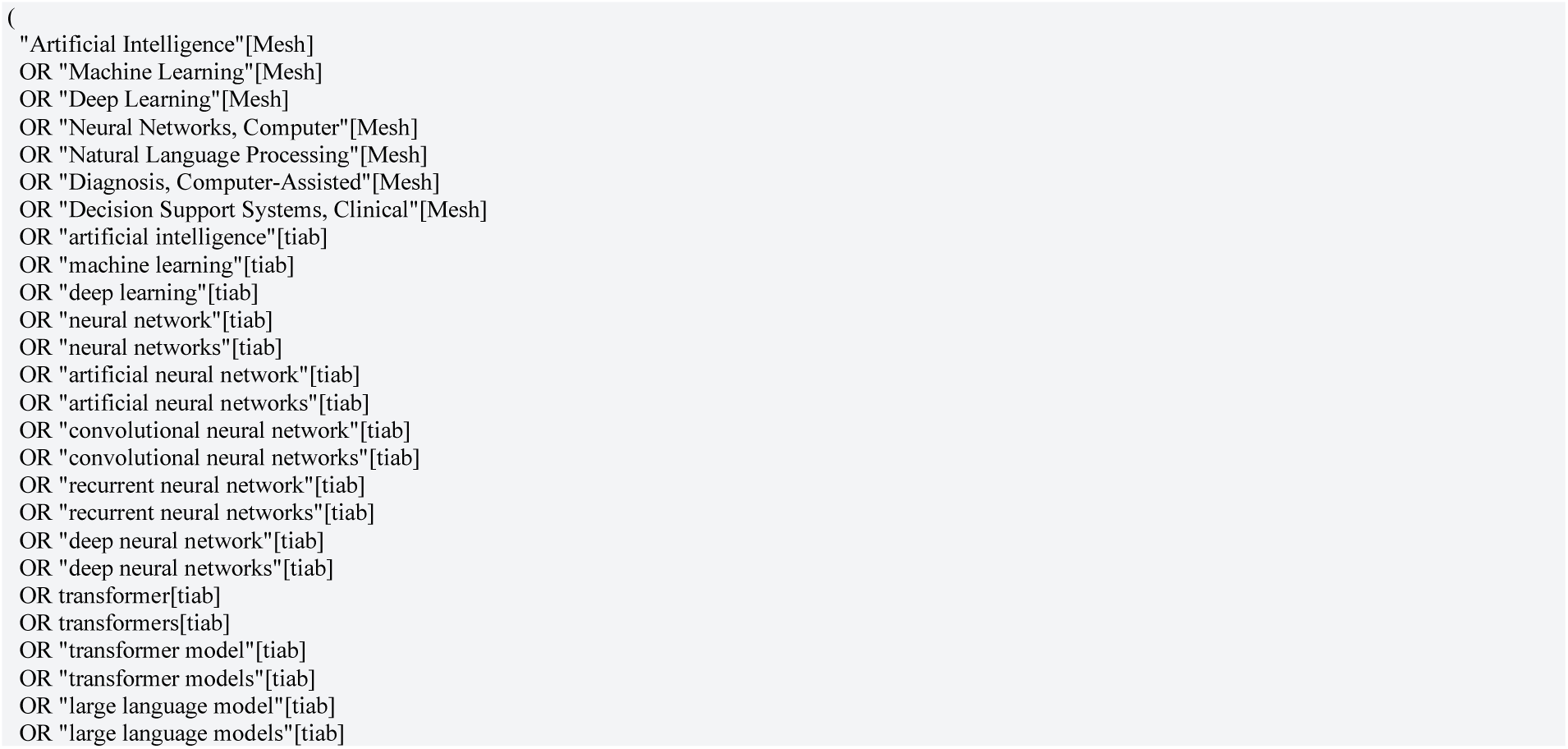
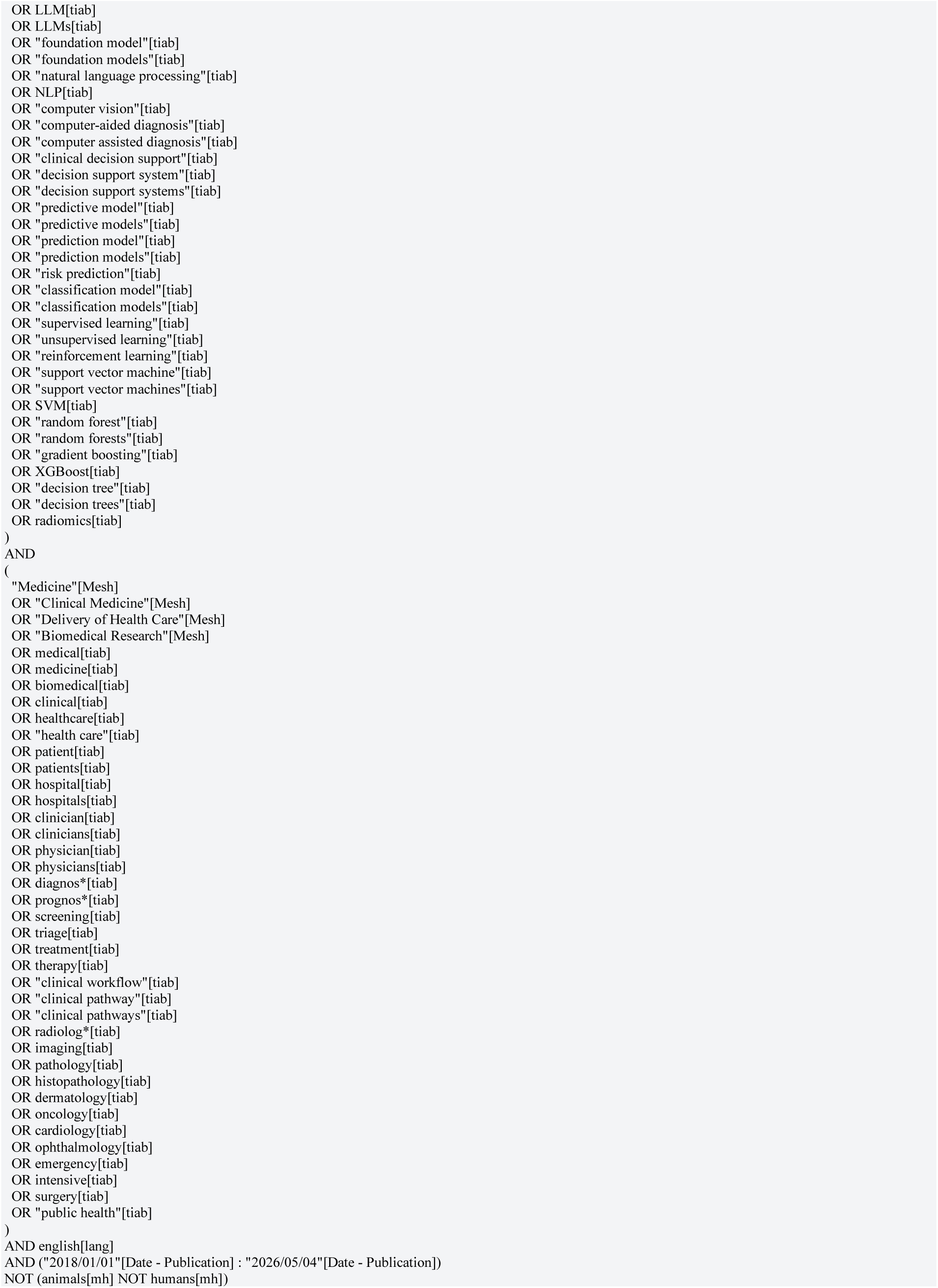

### 3. Publication handling

Reviews, editorials, commentaries, conceptual papers, and non-evaluable method papers that passed the search were labelled as class 5- not applicable.

### 4. Post-search heuristic AI screen

After retrieval, we retained records whose titles or abstracts contained at least one of the following AI terms. This screen narrowed a deliberately broad corpus before sampling and annotation.

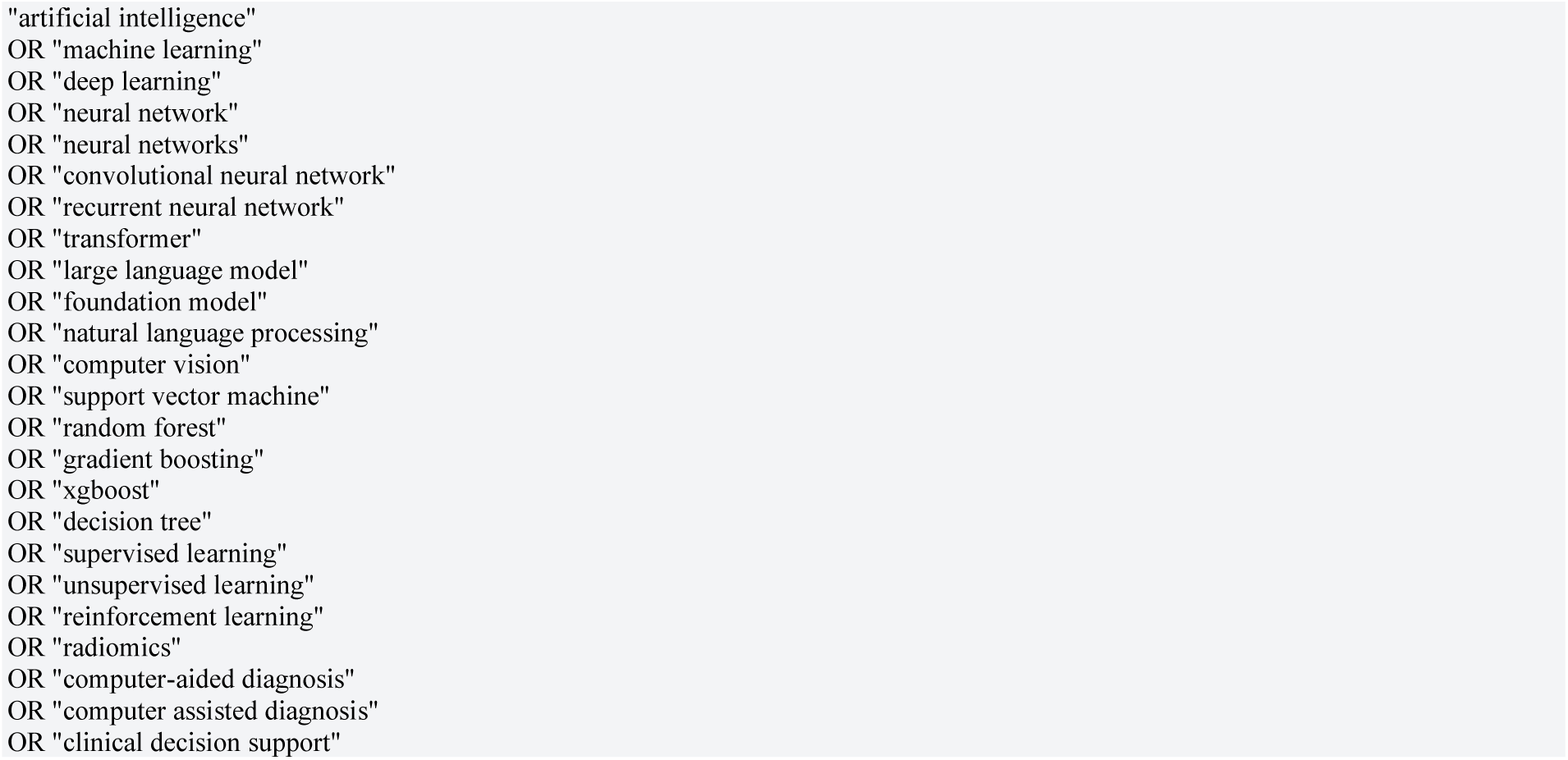

#### 4.1 R implementation of the heuristic screen

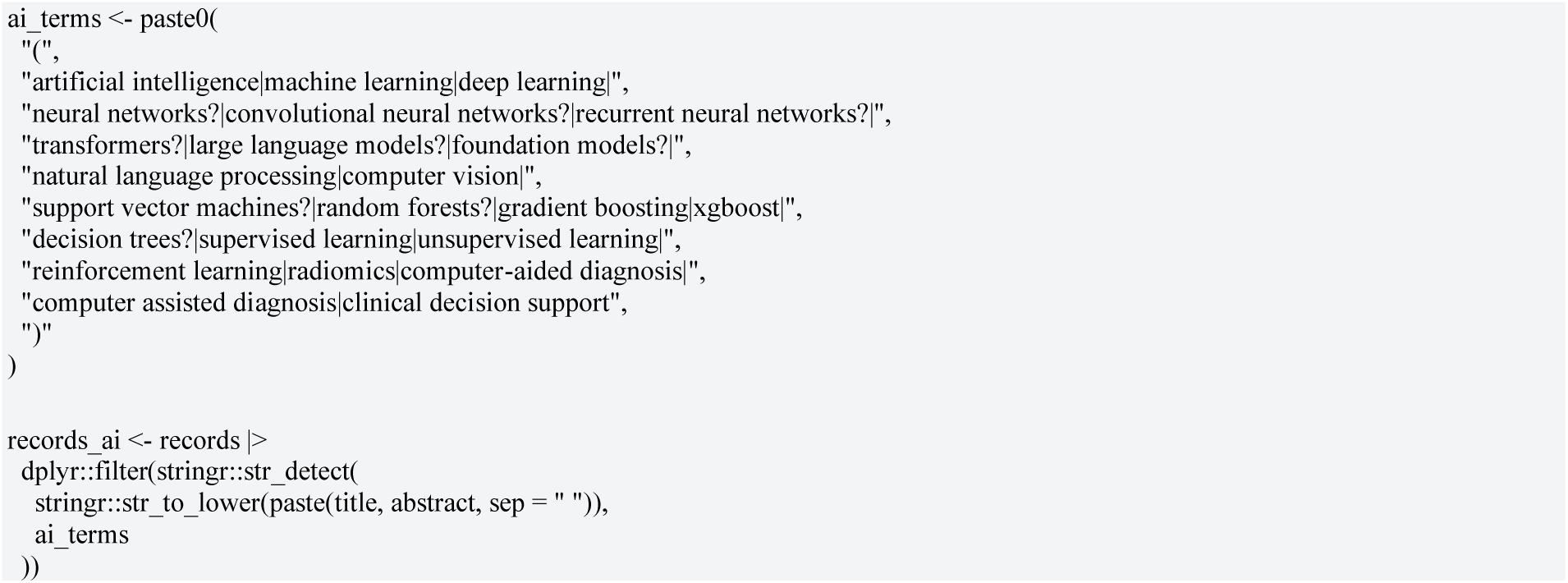

### 5. Minimum extraction and deduplication fields

| Field | Purpose |
| --- | --- |
| PMID | Primary unique identifier |
| DOI | Secondary identifier |
| Title | Near-duplicate detection |
| **Abstract** | Text used for classification |
| **Publication year** | Sampling balance and trend analysis |
| **MeSH terms** | Metadata and sensitivity analyses |

### 6. Out retrieval workflow

1. Ran the main PubMed broad corpus query.
2. Exported PMID, DOI, title, abstract, journal, publication year, publication type, and MeSH terms.
3. Removed records with no retrievable abstract if the model uses title/abstract text only.
4. Applied the post-search AI heuristic screen to the title and abstract.
5. Deduplicated by PMID, DOI, and normalised title hash; inspect near-duplicates with string distance.
6. Annotation set with year balancing from 2018 to 4 May 2026.
7. Double-annotated and reconciled maturity labels before model training.
8. Reserved-maturity-enrichment searches for class-balancing or sensitivity analyses, not for the primary prevalence estimate.

